# Food intake patterns, social determinants and emotions during COVID-19 confinement

**DOI:** 10.1101/2022.02.11.22270876

**Authors:** Maritza Rios, Jon Subinas, Celestina Delgado, Eliecer Torres, Amador Goodridge, Idalina Cubilla-Batista

**Author notes:** **CORRESPONDING AUTHORS:**, Postal Address: Aguadulce City, Coclé, Panama, Postal Address: Building 208, Ciudad del Saber, Panamá. Contributed equally to this work as co-first authors (Maritza Rios, Jon Subinas).

## Abstract

**Objective:** The COVID-19 pandemic was accompanied by varying movement restriction measures across populations worldwide. These restrictions altered daily activities at all levels, including food access and intake, as well as psychological feelings during lockdown. The main objective of the present study was to evaluate health, and nutrition behaviors during confinement during the first wave of the COVID-19 pandemic.

**Method:** We conducted a cross-sectional study using an online survey for data collection; a total of 1,561 surveys were validated.

**Results:** The majority of respondents were women (74.2%) between 18 and 49 years old. Among the respondents, 83.3% indicated a university education level, and 49.9% reported a monthly family income equal to or less than 1,000 USD. In addition, more than 50% self-reported overweight or obesity. Responses were analyzed using *k-means* algorithms to identify food intake patterns; we found three patterns: a healthy food intake pattern, a non-healthy food intake pattern and a mixed food intake pattern. The respondents with healthy food intake and non-healthy food intake patterns reported better socioeconomic conditions. Individuals classified as having mixed food intake patterns had lower incomes, less education and higher unemployment rates. Regarding emotions, we found that women experienced more negative emotions, such as fear, worry and anxiety, during the lockdown period.

**Conclusions:** Taken together, these results suggest that the mobility restriction measures imposed during the COVID-19 pandemic affected food intake patterns by exacerbating existing inequalities. We believe that directing resources towards strategies with the greatest positive impacts on public health remains key, especially in critical situations such as the COVID-19 pandemic.

## INTRODUCTION

At the end of 2019, SARS-Co-2 was identified in China as the virus causing coronavirus disease COVID-19. Due to the virus’ speed of expansion and severity, on March 11, 2020 the World Health Organization (WHO) declared a pandemic [1, 2]. The first case of COVID-19 in Panama was announced on March 8, 2020, and two weeks later, on March 24, Panamanian authorities declared a national lockdown with mobility restrictions according to gender and ID number [3]. These restrictions were by far the strictest in the Americas and were in place for nearly four months. Such a disruptive event has had a negative impact on the country’s economic, social and health conditions and augmented the long-standing social inequities among vulnerable groups [4]. In any pandemic, increases in food insecurity and job instability are expected, along with changes in daily life patterns and the appearance or exacerbation of sleep problems and other mental health issues; the COVID-19 pandemic was no exception [5-8].

Panama has experienced a heavy COVID-19 caseload resulting in a total of 737,689 cases and 7,909 deaths (February 2022). As a consequence of the virus and the extensive quarantines, the Isthmus experienced a 17.9% decrease in its Gross Domestic Product [9], together with an accelerated 20% increase in public debt and high levels of unemployment, which reached 18.5% in 2020. Unfortunately, in 2021 more than 130,000 residents in Panama fell elow the poverty line of $ 5.5 per day [4]. These changing economic conditions undoubtedly lead to changes in behavioral patterns.

Optimal dietary and lifestyle patterns are essential for preserving the body’s abilities to fight any infectious disease, in particular COVID-19 [10-12]. Adequate protein intake is required for antibody production, while low micronutrient levels favor viral infection, and high glycemic index meals promote an increased inflammatory response [13-17]. Therefore, a balanced diet characterized by high-quality proteins, micronutrient-dense foods and limited refined carbohydrates is recommended in the current pandemic situation.

In critical situations, such as a pandemic, altered living conditions can modify eating styles and influence other health behaviors, which may worsen malnourishment that existed before the arrival of SARS CoV-2 [18]. Quarantine is characterized by limitations to free circulation and changes in daily routines in the home and workplace, which may result in changes in emotional states, such as anxiety, or changes in physical activity and eating patterns [19-21]. This research aims to evaluate nutritional patterns in addition to health and social factors as measured during the COVID-19 lockdown among adults in Panama.

## MATERIALS AND METHODS

### Study design, location and population

The survey was targeted to Panamanian citizens and residents aged 18 years or above. The sampling frame for online research was non-probabilistic, that is, not all members of the population (N) had the same probability of participating due to the digital gap between those with and without access to communication technology. The majority of participants reported a university-level education and a monthly income above 2,000 USD. Another group of participants that reported lower educational levels or fewer resources was included in order to compensate for this bias. Efforts were also established in the online profile to survey residents living in rural provinces as well as indigenous comarcas. Our study was registered with the Ministerio de Salud of Panama (Registry No. 1,535) and was approved by the Comité Nacional de Bioética de la Investigación (CNBI, No. EC-CNBI-2020-0677).

### Surveys

The online surveys included questions related to sleep time, physical activity (PA), changes in emotions, alcohol consumption, use of nutritional supplements before and during the pandemic, criteria used for food acquisition, perceived quality of food intake and changes in the number of daily meals during the pandemic period. Food intake was measured using food frequency questions that focused on 20 food items from the Panamanian diet taken from the “Guías Alimentarias para Panamá” [22]. To evaluate changes in nutritional behavior, the survey included questions regarding food intake change before and during mobility restrictions were put in place. (see Supplementary data #1)

### Online participant recruitment

An online research strategy was implemented, and the survey was disseminated via different social channels, including high-volume social networks. This allowed the questionnaire to be answered by sufficiently diverse profiles. Several NGOs were contacted to disseminate the questionnaire among the most socioeconomically depressed areas. These strategies were complemented by sharing the questionnaire with personal contacts via WhatsApp and the use of online advertising on social networks. In addition, personnel from health centers in rural and indigenous areas were contacted to complete interviews in areas with lower network coverage and digital access. These strategies were aimed at increasing participation among respondents with low economic and educational resources, which is the population most difficult to reach via online forms.

### Face-to-face participant recruitment

64 In-person interviews were conducted at Health Ministry clinics. Registered nutritionists randomly selected visiting out-patients and applied the same survey using a mobile phone or PDA (Personal Digital Assistant). These responses were added to our survey database and analyzed together with those of the online respondents.

### Data collection and analysis

The online survey software Alchemer (www.alchemer.com, formerly SurveyGizmo, Louisville, CO, USA) was used to collect responses. The Alchemer application created reports in which the responses and profiles of the respondents were made visible in real time, allowing the research team to analyze information related to the data collection process. Reports were created to monitor information collection. A series of validation steps were followed, and the surveys determined to be incomplete were excluded from the final analysis. Data processing and analysis were completed using statistical packages SPSS (Social Package for Social Sciences) and STATA (15.1). We conducted categorical and bivariate analysis. We also applied the k-means clustering technique, or distance-based algorithm, in which we calculated distances from a mean to assign a point to a *cluster*. This allowed us to characterize food intake patterns among the study participants. Specifically, we used the self-reported changes in intake patterns for rice, tubercles (tubers), pulses, vegetables and green leafy vegetables, fruits, meat, sea products, chicken, pork, viscera (organ meats), eggs, dairy, fat from fruits and animals, fat from seeds, ultra-processed meat, fast food, sugary drinks, pastries and confectionaries (desserts or sweets), tea or coffee with sugar, and alcohol. Based on the k-means clustering method, three food intake patterns were defined as follows: -1 for lower consumption, 0 for same or nonconsumption and +1 for increased consumption as compared to before the lockdown restrictions. For example, a person that decreased their meat intake during lockdown was categorized into group -1. We used these three patterns to classify our study participants and analyzed these patterns according to their reported demographics, emotions and feelings, and other health behaviors during the confinement period.

All results are reported as percentages and frequencies for the entire sample and for each food intake pattern; a *chi* square test was used to analyze differences between the dietary patterns described. The variables related to emotions were evaluated using a bivariate analysis which included sex, age, educational level, income level and place of residence as classifying variables (see Suppl data).

## RESULTS

Between June 1^st^ and August 15^th^ 2020 we administered an online survey to a total of 2,475 participants over the age of 18. We also completed 64 face-to-face interviews. After data validation, a total of 1,561 surveys were included in the study. The majority of the participants were women (1,158 (74.2%)) between 18 and 49 years of age. A total of 1,211 (77.9%) participants reported living in Panama City and Panama Oeste, while 350 (22.1%) participants were living in other provinces and indigenous comarcas. Among the respondents, 1,297 (83.1%) reported a university education level, and 782 (50.1%) reported family incomes greater than 1,000 USD. Table 1 describes the participants’ demographic and socioeconomic characteristics. Regarding clinical conditions, 69 (4.4%) reported being positive for SARS-CoV-2 infection at the time of the survey.

**Table 1.** Sociodemographics characteristic of the study participants.

|  | **Frequencies N(%)**<br>**(N=1,561)** |
| --- | --- |
| **Age** |  |
| 18-29 years | 516(33.1) |
| 30-49 years | 756(48.4) |
| ≥ 50 years | 287(18.4) |
| Median Age | 35, IQR=63 |
| **Educational level** |  |
| University level | 1,297(83.1) |
| No University | 264(16.9) |
| **Montly family income** |  |
| ≥1000 USD | 782(50.10) |
| **Monthly food expenditure** |  |
| 0-100 USD | 263(16.8) |
| 101-200 USD | 356(22.8) |
| 201-300 USD | 355(22.7) |
| 301-400 USD | 265(17.0) |
| 401-500 USD | 155(9.9) |
| >500 USD | 167(10.7) |
| **Employment status** |  |
| Employed | 749(48.0) |
| Unemployed | 341(21.8) |
| Self employed | 192(12.3) |
| Student | 153(9.8) |
| Retired | 67(4.3) |
| Housekeepers | 59(3.8) |
| **Type of housing** |  |
| House | 1,124(72.0) |
| Condominiun or Apartament | 380(24.3) |
| Multifamiliar | 44(2.8) |
| Otro | 13(0.8) |
| **SARS-CoV2 infection status** |  |
| No history of infection | 1,492(95.6) |

Differential feelings and emotions were observed among the study participants. Both men and women reported feeling worried; however, the proportion of women who reported feelings of anxiety, fear and sadness was significantly higher. A significantly greater proportion of men reported experiencing mixed feelings, such as anger and calm. In general, both men and women reported experiencing positive feelings such as joy and calm less frequently (Figure 1).

**Figure 1.**
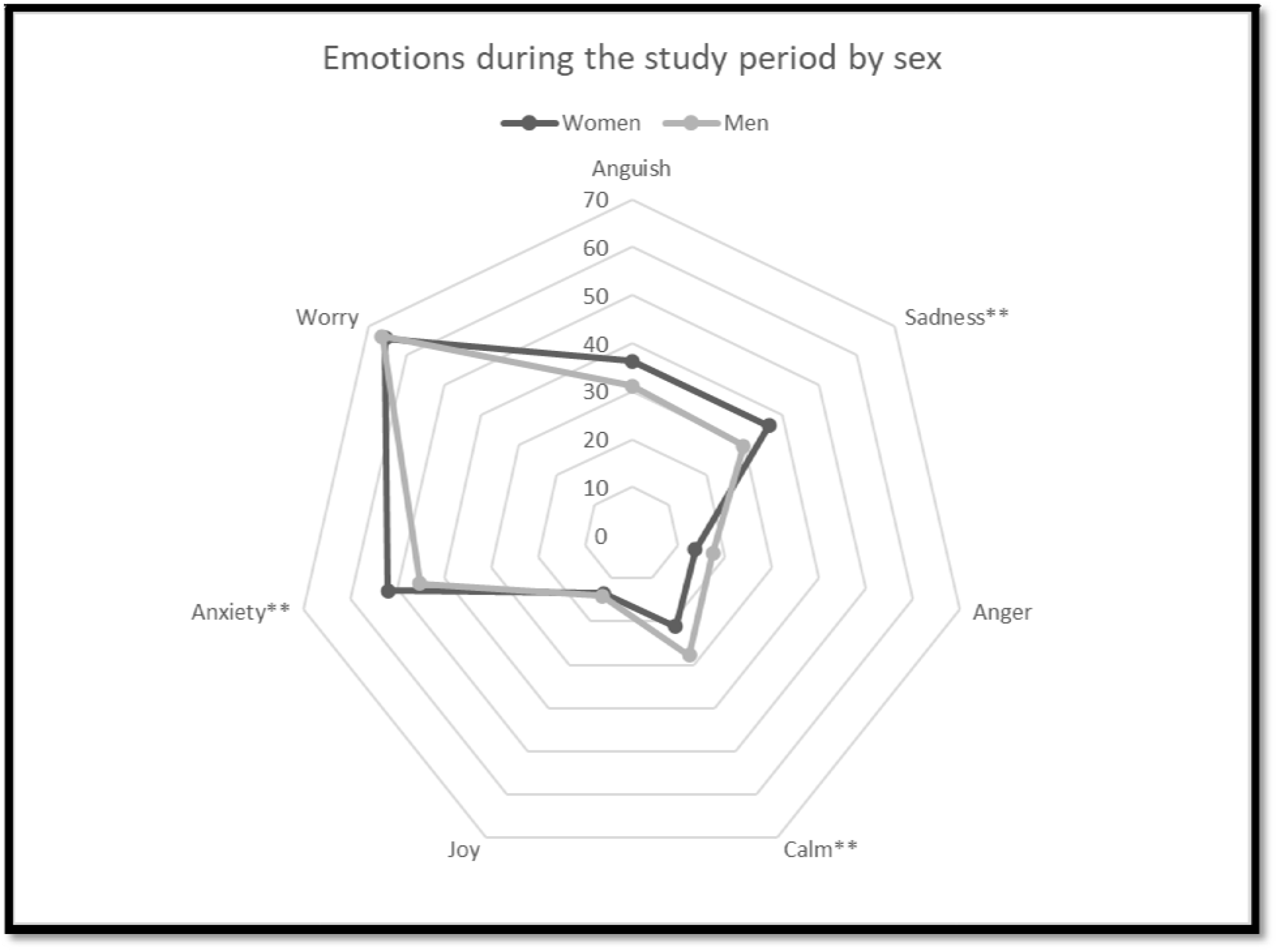
Emotions reported during the lockdown quarantine period in Panama by sex. Chi square test to compare women vs men. ** p<0.05

Overall, our study participants showed diverse food intake patterns. We classified our data set into three (3) food intake patterns using the *k-means* clustering method. Overall, from 40% to 60% of our study participants reported no changes in their food consumption, depending on the food item; however, in each food intake group, interpretable patterns were found among those who reported changes in their food consumption during the pandemic. Specifically, pattern 1 showed an increased intake of high-glycemic index foods or less healthy foods, such as bread, tortillas/empanadas, rice, processed meat, and tea or coffee with sugar, fast food, sugary drinks, and boxed cereals, pastries, confectionary (sweets and desserts) including ice cream, and alcohol during the study period. This same group reported a reduced intake of fruits and vegetables. In contrast, the pattern 2 participants showed an increased intake of fruits and vegetables, sea products, legumes, starchy vegetables, and fats from fruits and animals. The pattern 2 group demonstrated a reduced intake of bread, rice, tortillas/empanadas, processed meat, and sugary drinks, fast food and confectionary/pastries. Finally, the participants from pattern 3 showed a decreased intake of bread, vegetables, meat, and pork, but also tortilla/empanada, all types of cereals, legumes, fat from fruits and animals, and ultra processed meat. Interestingly, the pattern 3 group reported a consistent intake of rice, starchy vegetables, legumes, fruits, poultry, eggs, fast food, pastries, and confectionary items from the pre-pandemic to the pandemic time period.

The food pattern 2 group had the highest percentage of study participants with a university education; 9.4% of participants from this group reported lower education levels. In contrast, 26.3% of participants classified as food pattern 3 reported no university education. More than half of the respondents from the food pattern 1 and food pattern 2 groups reported begin employed, whereas unemployment was two times higher among respondents in the food pattern 3 group.

Nearly 65% of respondents from the food pattern 3 group had a monthly income less than or equal to 1,000 USD. On the other hand, about 60% of respondents from groups 1 and 2 reported a monthly income higher than 1,000 USD. In general, across the three groups, nearly 73% were living in a house during the pandemic, and 95% were not COVID-19 positive at the time of the survey.

Among the 762 participants who answered at least one question regarding emotions, a higher percentage of respondents from the pattern 2 group reported experiencing joy (18% (73)) and calm (30% (127)). In contrast, a higher percentage of respondents from pattern 1 experienced negative emotions, such as sadness, anguish, and worry. During the study period, 63% (263) of respondents from pattern 2 reported sleeping between 7 and 9 hours per night, whereas less than half of the participants from patterns 1 and 3 reported this (t test, p < 0.001). On the other hand, 70% of respondents from patterns 1 and 3 reported low physical activity, while 47% of pattern 2 participants reported sedentary behavior. Self-reported underlying chronic diseases were distributed similarly across the three patterns (*p >0*.*05. See Table 2))*. Almost half (49%) of pattern 1 participants reported that they were gaining weight during the pandemic. In contrast, 80% and 72% of the respondents from patterns 2 and 3, respectively, reported weight loss or no change in weight during the same period.

**Table. 2.**
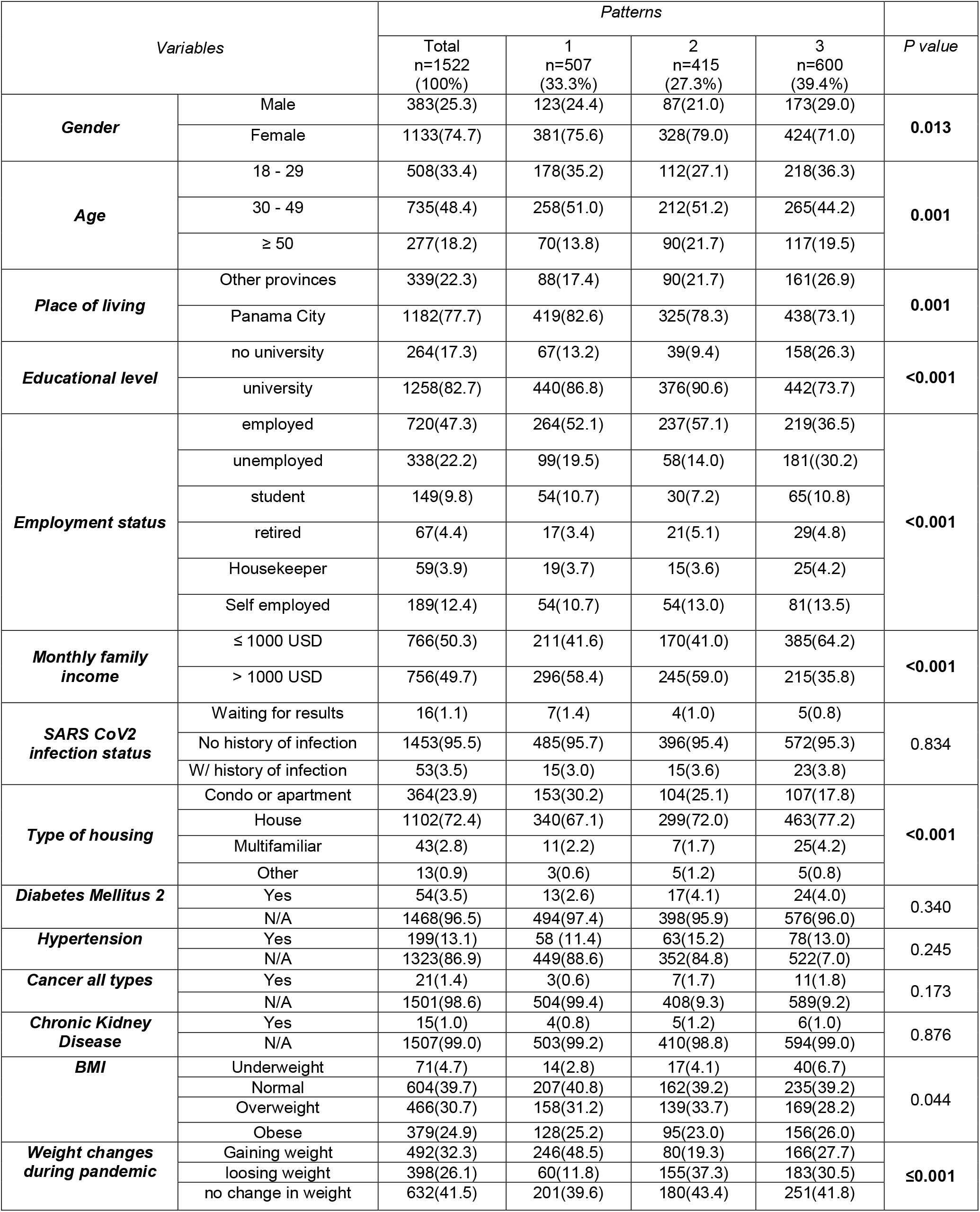
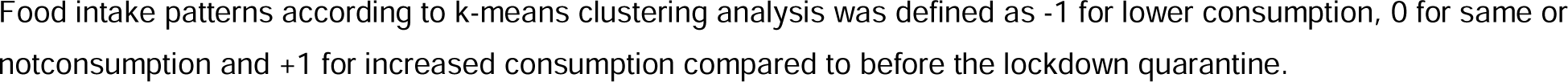
Characteristic of survey partipants for each dietary patterns.

**Table 3.**
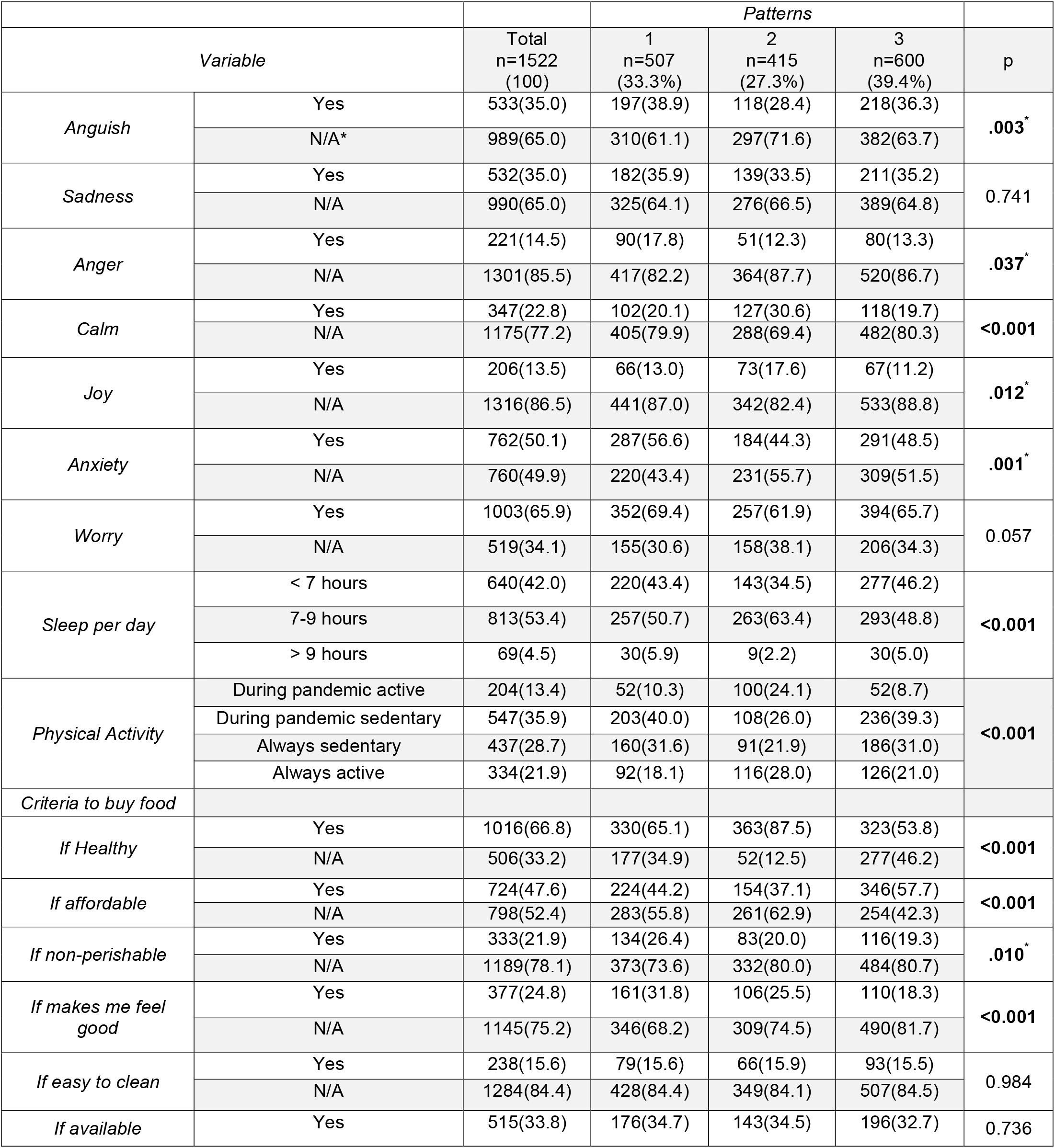
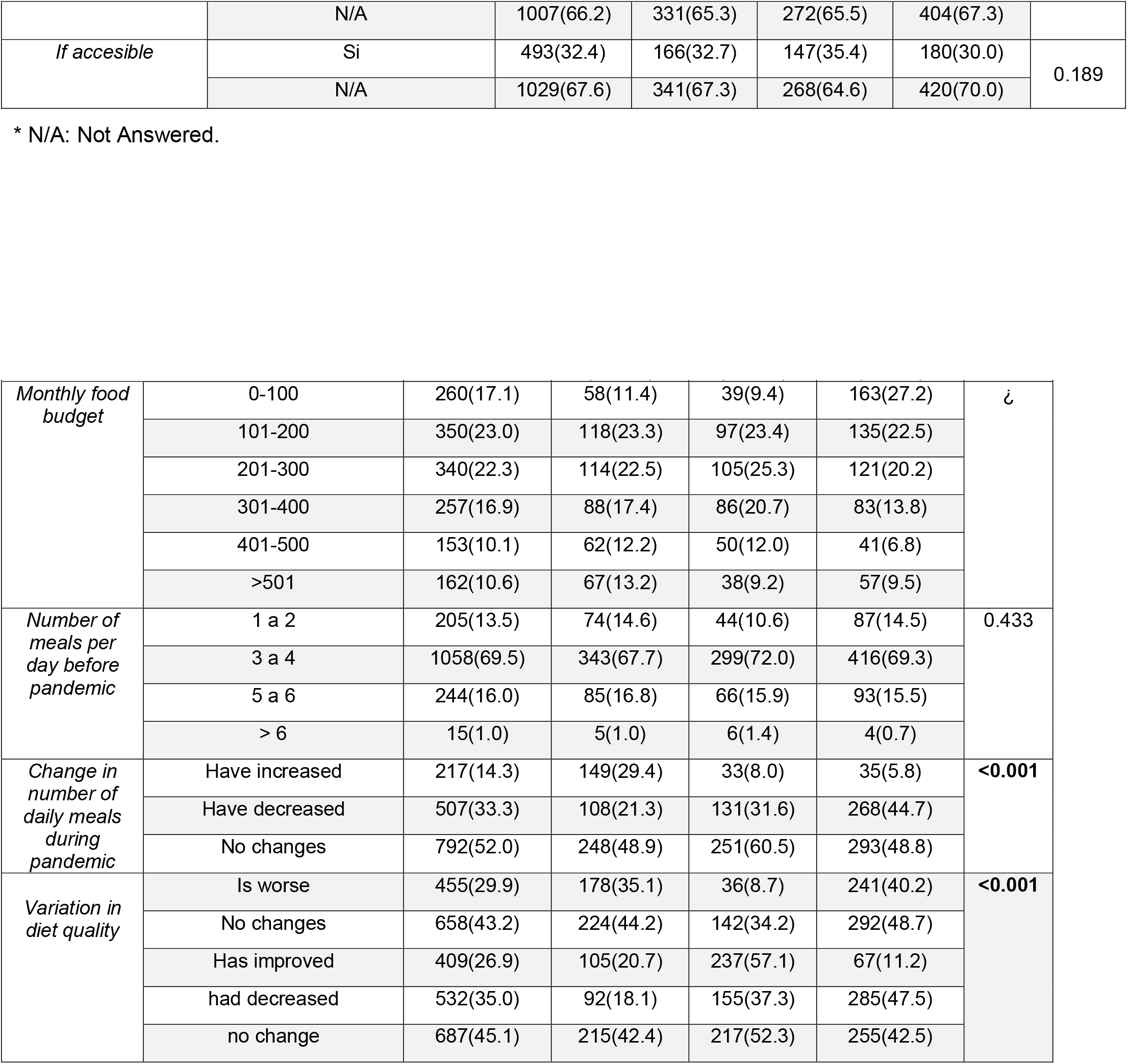
Health and food acquisition behaviors by each dietary pattern in Panama during COVID-19 lockdown quarantine.

**Table 4.**
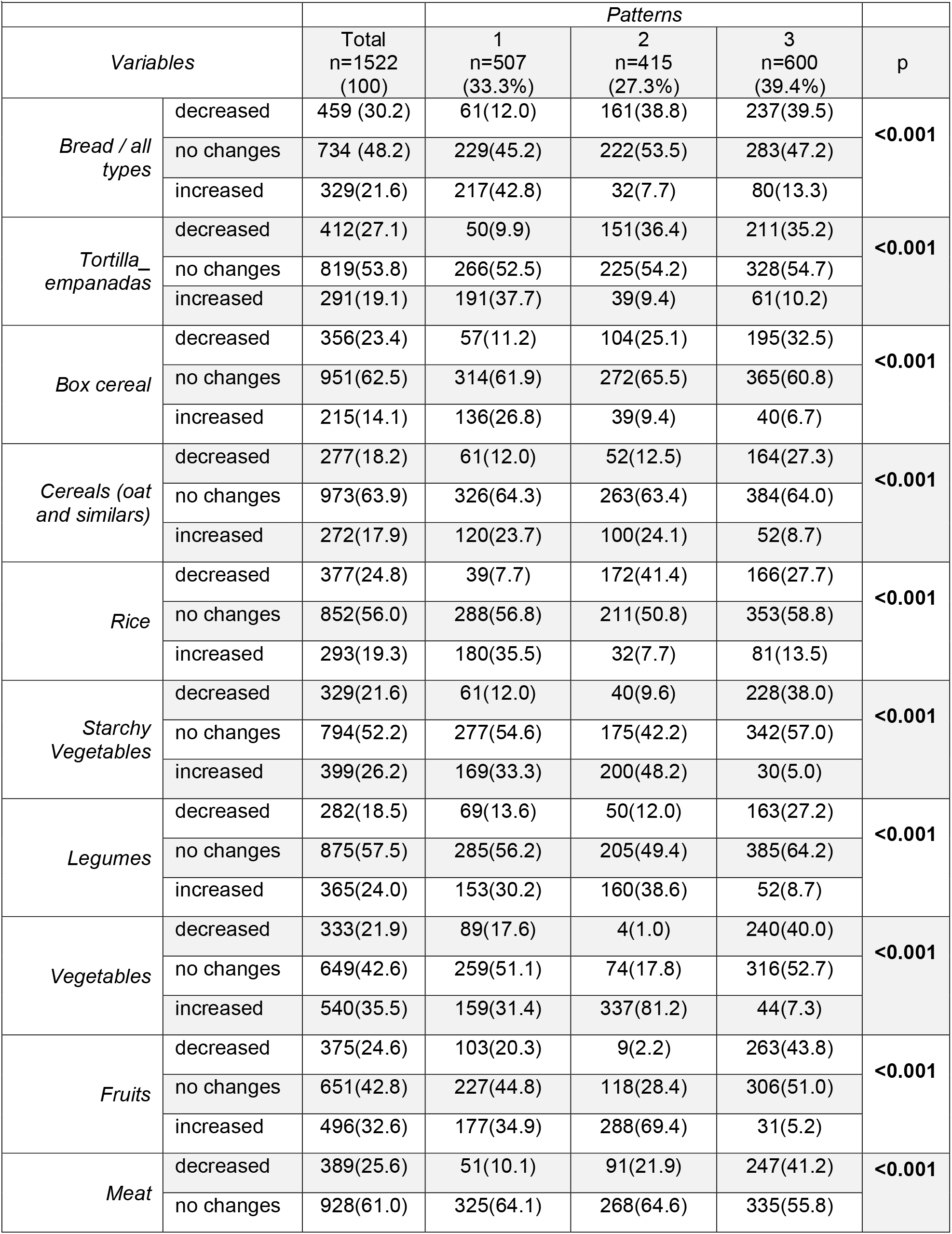
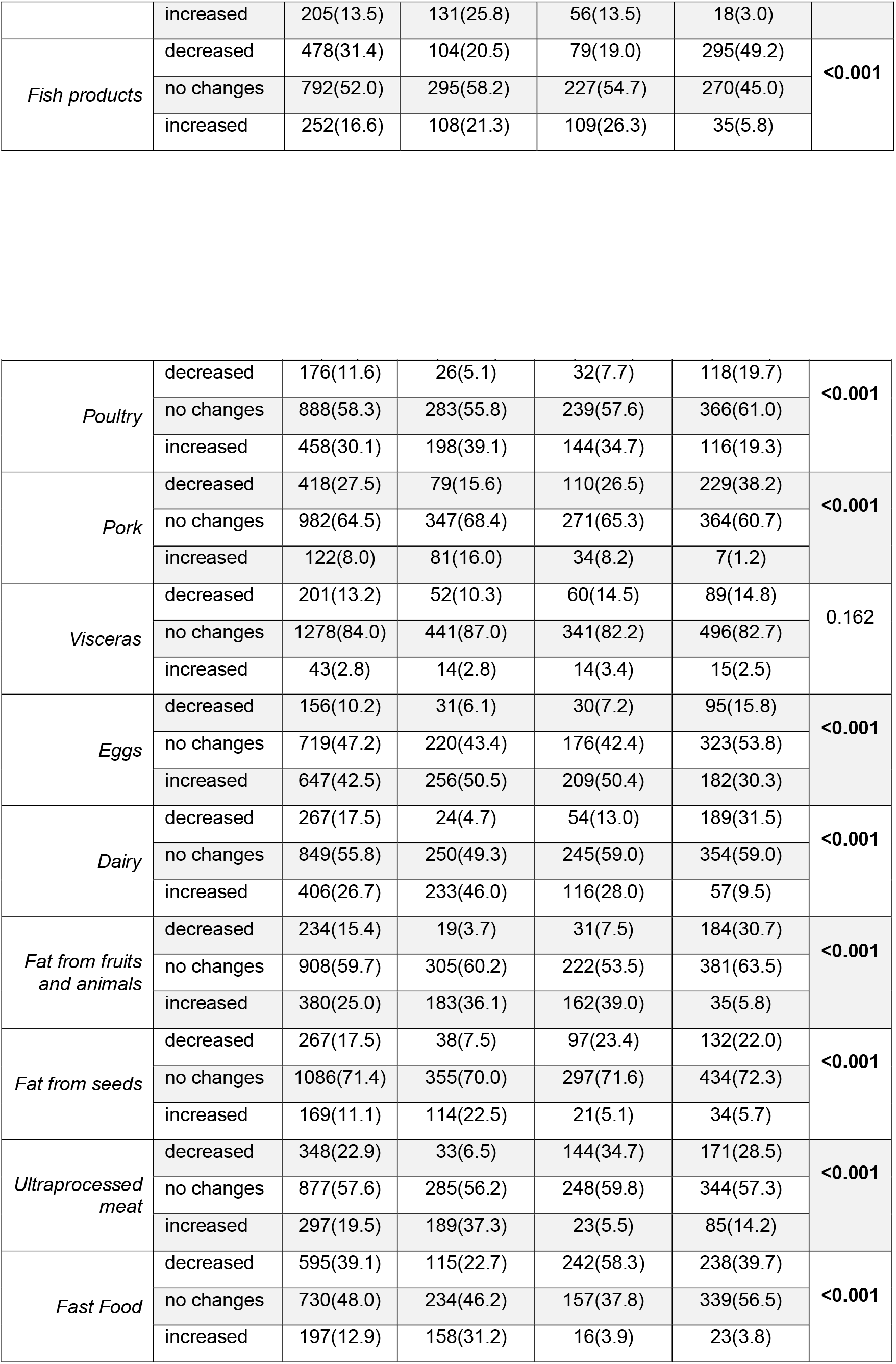
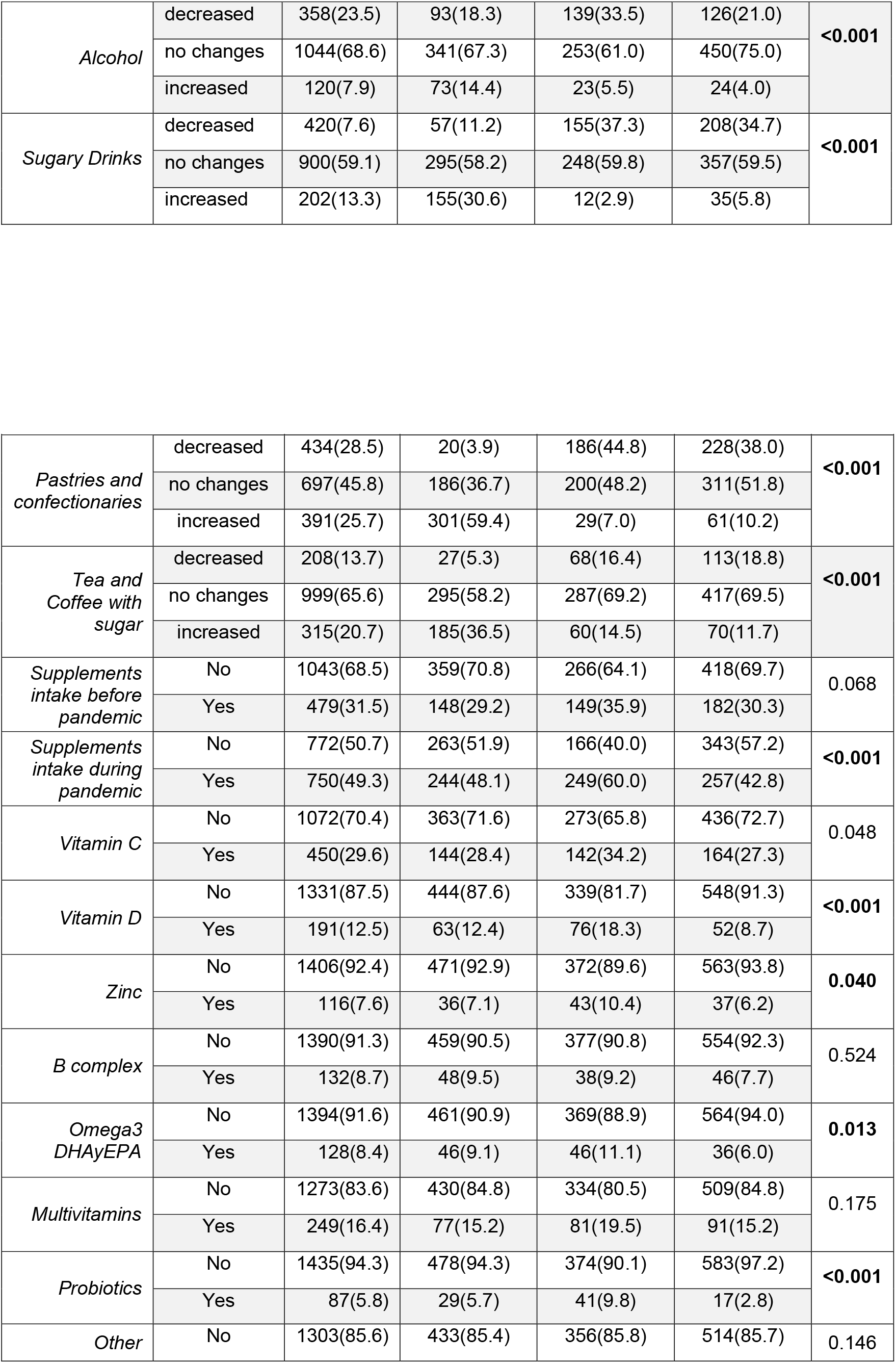
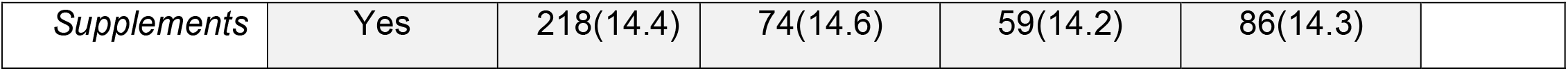
Food intake behavior by each dietary pattern.

We also found that nearly 88% of respondents from pattern 2 chose their food based on the fact that they were healthy and natural. More than 45% of pattern 1 and 3 respondents selected the type of food to purchase based on price. Finally, the highest percentage of participants who chose their food based on their emotions and the food’s health benefits as well as if the foods were nonperishable was from the pattern 1 group. About 70% of respondents from the pattern 3 group reported spending 300 USD or less per month on their family food needs.

Participants from pattern 2 reported better socioeconomic conditions. This group showed lower unemployment rates compared to groups 1 and 3. About 60% of the group 2 participants had a monthly income higher than 1,000 USD, while 35.8% of group 3 reached that monthly wage. Respondents from the pattern 2 group represent the highest percentage of participants with a university education, 4% higher than the pattern 1 group and 17% higher than the pattern three group. It is also interesting to highlight that the pattern 2 group reported a decreased consumption of poultry and eggs, which are typically more accessible protein sources.

Most of the participants across the three patterns (70%) reported eating 3 to 4 times a day before the pandemic; and for the pandemic period, 52% of individuals reported no changes in the number of meal times. The results demonstrated that the percentage of individuals who increased the number of meals consumed per day was three times higher among inviduals from the pattern 1 group compared to patterns 2 and 3. The survey included a self-evaluation of food intake quality during the pandemic; while 43% of all respondents perceived no changes in quality, 57% of respondents from the pattern 2 group reported an improvement in the quality of their food intake during the pandemic.

Regarding the use of vitamin supplements, across the three intake patterns, less than 35% reported supplement intake prior to the pandemic. In contrast, nearly 60% of participants from pattern 2 reported consuming vitamin supplements during the pandemic. The results demonstrated that about 40% of healthy food intake respondents had started using Vitamin C and D supplements. In general, a higher percentage of individuals from pattern 2 reported consuming Zinc, B complex, Omega 3, multivitamins, and probiotics.

## DISCUSSION AND CONCLUSIONS

Our study provides an overview of health and food intake behavior changes during the COVID-19 lockdown in Panama and the social and economic characteristics associated with each pattern identified. According to the results, respondents from pattern 1 have health and nutritional behaviors leaning toward what we called a ***non-healthy food intake pattern***; they exhibit a high consumption of high glycemic index foods, highly processed foods, and fewer fruits and vegetables. In contrast, participants from pattern 2 demonstrated health-promoting behaviors with a high consumption of foods with high micronutrient and fiber content, such as fruits, vegetables and legumes, and a remarkable pattern towards decreasing processed carbohydrates (white flour and refined sugar), and we interpreted this as a ***healthy food intake pattern***. The pattern 1 and 2 groups show similar levels of education, monthly income and employment status but nearly opposite health behaviors. The individuals from pattern 3 had a mixture of positive and negative behaviors but in general are characterized by a steady decrease in consumption of foods considered health promoters and health protective habits, such as physical activity and adequate sleep time, and we interpret this as a ***mixed food intake pattern***. For this last group, the number of meals consumed per day decreased during lockdown and half reported a monthly food expenditure equal to or less than 200 US, which does not meet the cost of the basic food basket in Panama [23]. This mixed food intake pattern is of concern because it was reported among 40% of our sample, and the group also showed economic limitations. The social characteristics of participants from patterns 1 and 2 vary from those belonging to the pattern 3 group, and these differences signal potential socioeconomic inequalities. Inequalities may explain non-healthy behaviors and decreased consumption of healthy food. On the other hand, the contrasting behaviors found in patterns 1 and 2 is a clear reminder that having resources does not always translate into positive decisions regarding food intake or health behaviors. Furthermore, other variables, such as self-perception of health status, social networks, and social and gender inequalities should be taken into account when it comes to formulating public health policies to promote healthy behaviors [24-26].

Recent studies have also confirmed the impact of low income and food insecurity on the nutritional behavior of family members [27, 28]. This is particularly troubling since Panama has experienced accelerated development during recent decades. Despite this progress in development, the country has not been able to reverse the high inequality rates and remains among the countries with the most unequally distributed wealth in the region [29]. This level of inequality persisted and was most likely increased during the COVID-19 pandemic.

Social and economic factors have an influence on individuals’ decisions regarding food consumption, while certain customs and local dietary practices are also involved. However, the impact of the COVID-19 crisis is different in countries where the public sector has built a powerful shield of social protection compared to countries where that shield has been much more fragile and high rates of inequality persist. In Panama, it is estimated that the Gini index will increase by 4 points, and relative poverty will increase by 3 points as a result of the pandemic [30].

In our study, the non-healthy and mixed food intake patterns, patterns 1 and 3, respectively, were characterized by other non-healthy behavior patterns, such as more sedentary lifestyle and more reported feelings of anxiety, worry and anguish. These results are similar to those found in other countries [31-33], where mobility restriction measures forced many to work from home and people could not exercise because gyms and even public parks were closed. According to the latest National Health Survey, ENSPA, 34% of the surveyed population was overweight before the pandemic [34]. The increase in sedentary lifestyle during quarantine will have a devastating consequence on the health of the population for which action should be taken. The Ministry of Health carried out campaigns to promote exercise at home using social networks for this purpose. However, for those in pattern 1, more aggressive campaigns should be designed in collaboration with local television stations to dedicate segments from broadcasting to exercise routines for all ages and promote decreased consumption of sugars, fats and oils as a way to prevent obesity and other chronic diseases [22].

Participants from the healthy food intake pattern are in a better social situation, have lower unemployment rates, higher incomes, and a higher level of education compared to the non-healthy and mixed food intake patterns. Also, we noted that the mixed intake group reported decreased consumption of poultry and eggs, which are typically more accessible protein sources, as well as Panamanian staples, such as tortillas, rice, and pulses. Taken together, our results support the notion that healthy food may not be accessible to the everybody. People may have the knowledge of what they should eat to preserve health, but without policies to address or reverse these socioeconomic disadvantages, food intake behaviors will not improve. The dramatic increase in unemployment and global economic recession have impacted every country [35]; our findings for Panama add to the regional evidence that specific efforts are needed to help the most vulnerable groups meet their minimum nutritional requirements to maintain health.

In social terms, the results of our study also reveal two different patterns of inequality related to eating behaviors. We believe that people from the mixed pattern, who are consuming less food, have a lower educational level, lower incomes, and a higher unemployment rate, are the most affected by the pandemic. This population is accumulating more vulnerabilities and has been forced to reduce their food consumption. As a consequence, they are prone to a severe deterioration of health related not only to eating patterns but to emotions, physical exercise, and sleep. In the mixed pattern group, we noted a subgroup who have maintained a “constant” diet and have a lower education level, low income and high unemployment. In this case, the data may be describing a chronic inequity in which the profiles that accumulate more vulnerabilities maintain their pre-pandemic diet because they do not have the means to make changes. This group spends less than 300 USD/month on food. According to WHO data, there is an increase in chronic diseases sequelae, setting up a scenario in which COVID-19 disease has the potential to affect the public health system [2]. Our findings suggest there is a need for a holistic approach to equip the population with the tools and resources to make healthier decisions, even in critical situations.

This study has limitations. The use of an online survey and self-reported behaviors carries bias; in order to minimize this bias, we did not ask for identification data, and the survey was built using software with data check options. The research team recognizes the shortcomings of identifying dietary patterns based on a reduced number of food items, but those included in this survey have been identified as the most consumed in Panama and are mentioned in the GABAS (from the Spanish: *Guías Alimentarias Basadas en Alimentos*). Previous studies have used similar items to identify food intake patterns [32, 36-38]. The sample size and the high participation rate were the study’s main strengths, constituting, to our knowledge, the first population-based study on dietary patterns in cooperation with the social sciences.

## Conclusions

This study suggests a high intake of cheaper protein sources, such as chicken, eggs and dairy during the pandemic period, which has been described elsewhere. However, a large proportion of the survey participants reported dietary consumption patterns similar to the pre-pandemic period. Healthy lifestyle and food intake habits were observed in the groups with the highest income and educational levels. Promoting higher intakes of vegetables and seafood to prevent deficiencies in omega 3 (DHA and EPA), fiber and micronutrients may reduce morbidity from disease burden experienced during the pandemic.

To improve this scenario we recommend protecting, supporting, and helping to maintain healthier eating patterns using different strategies. First, educating consumers on the disadvantages of consuming starchy carbohydrates, such as highly refined cereals if these have become popular in the area, and advocating for the consumption of varied starchy carbohydrates, such as cassava, plantain, potato, yam, or otoe (vegetables). Second, encouraging low-income families to use locally available food and, if possible, cultivate the local land. Third, take the necessary steps to introduce good eating practices in local schools and other public institutions, for instance, by improving local Dietary Guidelines based on the population and their healthy dietary patterns. In turn, authorities must adopt initiatives to improve the population’s access to food: developing public policies that facilitate and promote an affordable price for basic food products, such as vegetables, fruits, proteins and dairy products; implementing public policies that strengthen the agro-Panamanian market, making products affordable for different socioeconomic profiles; and generating local trade that results in a further development of the national market. The government should strengthen national institutions and public policies to increase the Panamanian economy’s resilience in response to fluctuations in commodity prices and volatility in capital flows. These data point to several indications that healthy foods are not accessible to the entire population. Awareness and education campaigns play an important role, but eating habits will not improve without policies designed to improve healthy food access.

## Data Availability

All data produced in the present study are available upon reasonable request to the authors

https://youtu.be/Z2X306wc-JU

## ACKNOWLEDGEMENTS

We thank the Secretaria Nacional de Ciencia, Tecnología e Innovación (SENACYT) and the Sistema Nacional de Investigadores de Panamá (**SNI**). Also, we thank to these nutritionists from primary care level from Ministerio de Salud de Panamá including Licda Maydelaine Rodriguez, Lic. Crystal Hernández, Lic. Marymar Salazar, Lic. Edgary Patricia Vega Rojas, Lic. Julissa Lidibeth Navarro Carrasco. The authors also thank Dr. Tania Herrera and Dr. Fernando Diaz from Centro de Investigaciones Pacifica Salud for their valuable comments. Finally, the authors thank Anne Williams and Colleen Goodridge for critically reviewing this manuscript.

## FIGURES & TABLES

**Suplementary #1** - las 24 pregurntas.

