## Supplementary data #1 for "Food intake patterns, social determinants and emotions during COVID-19 confinement"

##### SUPPLEMENTARY INFORMATION #1

**Survey Questionnaire**

**Are you an adult citizen, Panamanian or resident in Panama?**

### ( ) Yes

## ( ) No

**GENERAL**

**1)What is your gender?***

#### ( ) Female

#### ( ) Male

#### ( ) Other

##### 2) What is your age?

##### 3) What is your province* District* County

##### 4) What is your educational level?

##### ( ) Elementary or basic

##### ( ) Secondary (middle)

##### ( ) University

##### ( ) None

**5) What is your current employment status?**

( ) Salaried

( ) Self-employed

( ) Retired/Pensioned

( ) Unemployed/Not working

( ) I work as a domestic worker

( ) Student

### 6) What is the approximate monthly household income*?

### ( ) 0 - 400 dollars (usd)

### ( ) 401 - 1000 dollars (usd)

### ( ) 1001 - 2000 dollars (usd)

### ( ) More than 2000 dollars (usd)

##### Pandemic Related Data

##### 7) Select the place where you live during this pandemic*.

##### ( ) House

##### ( ) Apartment in building

##### ( ) Room in a tenement or multi-family dwelling

##### ( ) Other:

### 8) Do you have a confirmed diagnosis of coronavirus? *

### ( ) Yes

## ( ) No

### ( ) I am waiting for the test result.

##### Data related to your health

##### 9) What is your approximate height?________________________________________

##### 10) What is your approximate weight?*_________________________________________________

##### 11) During the pandemic, have you:*

##### ( ) gained/are gaining weight

##### ( ) lost/ are losing weight

##### ( ) maintained/are maintaining your weight

##### 12) Which of the following feelings are you experiencing most intensely in the last few days (you can check more than one)*?

##### [ ] Fear/anxiety

##### [ ] Sadness

##### [ ] Anger

##### [ ] Calmness

##### [ ] Joy

##### [ ] Anxiety

##### [ ] Worry

##### [ ] None of the above

**13) Select below the disease(s) of which you have been diagnosed (more than one can be checked) ***

[ ] Diabetes type II (high blood sugar)

[ ] Arterial hypertension (high blood pressure)

[ ] Obesity/Overweight

[ ] Cancer

[ ] Kidney or renal disease

[ ] None

[ ] Other: _________________________________________________

#### 14) Select the activity related to body mobility (physical activity) that best fits your situation*.

#### ( ) I was sedentary before and during the pandemic -1

#### ( ) I was sedentary before the start of the pandemic and now I am physically active. 1

#### ( ) I was physically active before the beginning of the pandemic and have stopped. -1

#### ( ) I have always been physically active and continue to be physically active. 0

##### 15) How many hours of sleep are you currently getting per day?*_______________________

##### 16) What criteria do you take into account when choosing food during shopping in this pandemic (you can select more than one)? *

##### [ ]Health: I try to choose more natural and healthier foods.

##### [ ]Cost: I choose by price and buy the cheapest.

##### [ ] Food shelf life: I choose non-perishable foods (cans, bottles, vacuum-packed, frozen).

##### [] Emotional well-being: foods that give me a sense of good mood and emotional balance.

##### [ ] Packaging: I choose foods with packaging that can be sanitized at home.

##### [ ] Availability: I buy foods that I find available.

##### [ ] Accessibility: I buy the foods that are sold closest to my home

### 17) Of the total family income. How much money do you use for food expenses ( groceries) on a monthly basis? *

### ( ) 0-100 usd

### ( ) 101-200 usd

### ( ) 201-300 usd

### ( ) 301-400 usd

### ( ) 401-500 usd

### ( ) more than 500 usd

##### FOOD CONSUMPTION (beverages, food)

##### 18) What is the number of meals you usually ate per day BEFORE the pandemic (include things you ate between meals). *

#### ( ) 1 a 2

#### ( ) 3 a 4

#### ( ) 5 a 6

##### ( ) More than 6

##### 19) In general, do you think your diet during the pandemic*?

##### ( ) Has improved ( ) Has gotten worse ( ) Has stayed the same

##### 20) During this pandemic

|  | **Change (Variation)** |
| --- | --- |
| Number of meals per day : | Increased  Decreased  Has remained the same |
| The amount of food ingested per day: | Increased  Decreased  Has remained the same |
| Portion size in food consumption has: | Increased  Decreased  Has remained the same |

##### 21) En relación a la siguiente lista de alimentos evalue cual ha sido su comportamiento de consumo *

|  | **No consumption*** | **Equal consumption*** | **Consumption LESS than before the pandemic*** | **Consumption MORE than before the pandemic*** |
| --- | --- | --- | --- | --- |
| **Consumption report** | ( ) | ( ) | ( ) | ( ) |
| Bread (mold, flute, others) | ( ) | ( ) | ( ) | ( ) |
| Tortilla or empanada (fried or grilled) | ( ) | ( ) | ( ) | ( ) |
| boxed cereal | ( ) | ( ) | ( ) | ( ) |
| Cereals (oats, corn-starch) | ( ) | ( ) | ( ) | ( ) |
| Rice | ( ) | ( ) | ( ) | ( ) |
| Starchy vegetables (cassava, yams, potatoes, etc.) | ( ) | ( ) | ( ) | ( ) |
| Legumes (lentils, beans, beans, chickpeas and similar) | ( ) | ( ) | ( ) | ( ) |
| Vegetables (non-starchy vegetables) | ( ) | ( ) | ( ) | ( ) |
| Fruits | ( ) | ( ) | ( ) | ( ) |
| Meats | ( ) | ( ) | ( ) | ( ) |
| Fish/seafood (fresh and canned) | ( ) | ( ) | ( ) | ( ) |
| Chicken (any part of the chicken) | ( ) | ( ) | ( ) | ( ) |
| Pork (any part) | ( ) | ( ) | ( ) | ( ) |
| Viscera (liver, kidney, tripe) | ( ) | ( ) | ( ) | ( ) |
| Eggs (fried, boiled, scrambled) | ( ) | ( ) | ( ) | ( ) |
| Dairy products (carton milk, evaporated milk, powdered milk, yogurt) | ( ) | ( ) | ( ) | ( ) |
| Fresh food | ( ) | ( ) | ( ) | ( ) |
| Fats 1 (avocado, extra virgin olive oil, coconut oil, butter, etc.)) | ( ) | ( ) | ( ) | ( ) |
| Fats 2 Hydrogenated or industrialized (vegetable oils such as sunflower oil, soybean oil, canola oil, corn oil, margarine) | ( ) | ( ) | ( ) | ( ) |
| Frozen ingredients | ( ) | ( ) | ( ) | ( ) |
| Ultra-processed meat foods (sausages, bologna, canned hams, nuggets, etc.) | ( ) | ( ) | ( ) | ( ) |
| Consumes foods such as pizza, hamburgers, French fries delivered to your home or homemade fries | ( ) | ( ) | ( ) | ( ) |
| Liquor in any presentation (beer, wine, or similar) | ( ) | ( ) | ( ) | ( ) |
| Beverages such as sodas, commercial nectars or sweetened juices | ( ) | ( ) | ( ) | ( ) |
| Sweetened products (cookies, pastries, ice cream, gelatine) | ( ) | ( ) | ( ) | ( ) |
| Tea, coffee or creams with sugar | ( ) | ( ) | ( ) | ( ) |

**22) Were you taking supplements BEFORE the pandemic***

( ) Yes

( ) No

**23) What supplements did you start consuming DURING the pandemic?**

[ ] None

[ ] Vitamin C

[ ] Vitamin D

[ ] Mineral Zinc

[ ] B complex

[ ] Omega 3 (DHA and EPA)

[ ] Multivitamins

[ ] Probiotics (bacteria in capsules)

[ ] Others
